# Speech-in-Noise Difficulties in Aminoglycoside Ototoxicity Reflects Combined Afferent and Efferent Dysfunction

**DOI:** 10.64898/2026.03.23.26348719

**Authors:** Lina Motlagh-Zadeh, Diala Izhiman, Chelsea M. Blankenship, Hebatalla Hamza, David R. Moore, Dawn Konrad Martin, Angela Garinis, Patrick Feeney, Lisa L. Hunter

## Abstract

**Objectives:** Patients with Cystic fibrosis (CF) often receive aminoglycosides (AGs) to manage recurrent pulmonary infections, placing them at risk for ototoxicity. Chronic AG use can lead to complex cochlear damage affecting inner and outer hair cells, the stria vascularis, and spiral ganglion neurons. The greatest damage is typically in the basal cochlear region, which encodes high-frequency hearing, with additional involvement of more apical regions. While extended-high-frequency (EHF) hearing loss (EHFHL; 9-16 kHz) is often the earliest sign of AG ototoxicity, speech in noise (SiN) effects are rarely studied. Our overall hypothesis is that SiN perception difficulties in individuals with CF, treated with AGs, are related to combined cochlear and neural damage, primarily in the EHF range but also in the standard frequency (SF; 0.25-8 kHz) range. Three mechanisms that <u>contribute</u> to SiN perception were evaluated in children and young adults: 1) a primary effect of reduced EHF sensitivity, measured by pure-tone audiometry (PTA) and transient-evoked otoacoustic emissions (TEOAEs); 2) a secondary effect of subclinical damage in the SF range, measured by PTA and TEOAEs; and 3) additional neural effects, measured by middle ear muscle reflex (MEMR) threshold (afferent) and growth functions (efferent).

**Design:** A total of 185 participants were enrolled; 101 individuals with CF treated with intravenous AGs and 84 age and sex-matched Controls without hearing concerns or CF. Assessments included EHF and SF PTA; the Bamford-Kowal-Bench (BKB)-SIN test for SiN perception; double-evoked TEOAEs with chirp stimuli from 0.71 to 14.7 kHz; and ipsilateral and contralateral wideband MEMR thresholds and growth functions using broadband stimuli.

**Results:** Reduced sensitivity at EHFs (PTA, TEOAEs) was not associated with impaired SiN perception in the CF group. SF hearing, regardless of EHF status, was the primary predictor of SiN performance in the CF group. Increased MEMR growth was also significantly associated with poorer SiN in the CF group.

**Conclusions:** In CF, impaired SiN perception was primarily predicted by SF hearing impairment, with additional involvement of the efferent auditory pathway through increased MEMR growth. These results build on prior evidence for efferent neural effects due to ototoxic exposures, supporting both sensory (afferent) and neural (efferent) mechanisms that contribute to listening difficulties in CF. Thus, preventive and intervention strategies should consider these combined mechanisms in people with AG ototoxicity to address their SiN problems.

## Introduction

### Aminoglycoside Ototoxicity in Cystic Fibrosis Patients in Relation to Extended High-Frequency Hearing Loss

Cystic fibrosis (CF) is a life-threatening inherited disorder, most common in Caucasians, with a prevalence of approximately 1 in 3000 to 4000 live births (Sanders et al, 2016; Kreicher et al., 2018). It results in chronic pulmonary infections, which are treated with intravenous aminoglycoside antibiotics (AGs) when oral or inhaled antibiotics are not effective. Tobramycin is most commonly used, but amikacin and gentamicin, as well as glycopeptide antibiotics such as vancomycin, may also be administered (Barclay et al., 1996; Becker & Cooper, 2013; Jiang et al., 2017; O’Sullivan et al., 2017). AGs have contributed substantially to improved life expectancy in patients with CF, particularly before the introduction of cystic fibrosis transmembrane conductance regulator (CFTR) modulator therapies. According to the Cystic Fibrosis Foundation Patient Registry, the predicted median survival increased from 30 to 37.4 years between 1997 and 2008. More recently, the 2024 CF Foundation Registry reports that over half of patients receiving CFTR modulators now have projected survival into their 40s. These advances in longer lifespans among people living with CF underscore that quality of life, including preserving hearing function and treating significant dysfunction, is an important goal.

The cochleotoxic effects of AGs are well known to be associated with sensorineural hearing loss that begins with outer hair cell damage in the basal turn of the cochlea. AG antibiotics induce oxidative stress, which in turn causes hair cell loss in the basal cochlea regions responsible for encoding high-frequency sounds (Kros & Steyger, 2019; Campbell & Le Prell, 2018). Histopathological analyses of human (Matz et al., 1965) and animal (McDowell, 1986; Tange et al., 1982) temporal bones demonstrate that AG treatment leads to greater loss of outer hair cells (OHCs) in the basal high-frequency region of the cochlea, with less impact on OHCs in the apical low-frequency region. Ishikawa et al. (2019) also showed that low doses of AGs reduced the number of inner hair cell (IHC) synaptic ribbons in the basal turn of the cochlea, while OHCs remained unaffected. When AGs were applied at higher concentrations, OHCs were affected primarily in the basal regions of the cochlea.

These findings underscore the importance of extended high-frequency (EHF; > 8 kHz, basal cochlea) hearing assessment in ototoxicity monitoring in CF patients receiving AG treatment. Multiple studies showed greater EHF hearing loss (EHFHL) in CF patients than expected for age, despite normal standard frequency (SF; 0.25-8 kHz) audiograms (Blankenship et al., 2021; Garinis et al. 2021). Konrad-Martin et al. (2005) found that 30% of patients developed significant EHFHL before any noticeable changes in the SF audiometric range. Fausti et al. (1994) reported that 62.5% of ears with a decrease in pure-tone hearing sensitivity related to AG treatment have initial hearing loss solely in the high-frequency range. Although there has been extensive research into the pure-tone effects of AG treatment, the functional consequences for speech perception in noise and everyday communication function remain unclear.

### The Role of EHF Hearing in Speech Perception in Noise

Understanding speech in noisy, reverberant environments is the most common hearing complaint patients report, yet it is a poorly understood problem that varies significantly among individuals. Emerging research suggests that EHF hearing, beyond the SF range typically tested in the standard clinical audiogram, plays a significant role in speech-in-noise (SiN) perception (reviewed in Hunter et al., 2020), challenging previous evidence that speech perception relies only on frequencies below 7 kHz (Crandall & MacKenzie, 1922; Fletcher & Galt, 1950; Fletcher & Steinberg, 1930; Monson et al., 2014). For example, Best et al. (2005) demonstrated that individuals with better EHF hearing thresholds performed better on SiN tasks, highlighting the role of these higher frequencies in understanding speech in challenging environments.

Similarly, Trine and Monson (2020) reported that enhanced EHF hearing correlates with improved speech recognition in complex auditory settings, further supporting the importance of these frequencies for speech perception. Motlagh Zadeh et al. (2019) found that EHF hearing is important for distinguishing speech from competing sounds, emphasizing its role in preserving speech intelligibility in challenging listening conditions. Moreover, Flaherty et al. (2021) highlighted that EHF hearing was beneficial for speech recognition in complex listening situations for children as young as 5 years old. Collectively, these findings underscore the important role of EHF hearing in both adults and children and challenge the traditional view that speech perception only relies on lower frequencies.

Exploration of the underlying mechanisms connecting EHF hearing and SiN perception has recently attracted considerable interest, resulting in various hypotheses and mixed results. Some researchers suggest that the primary effect of EHFHL on SiN perception is due to reduced EHF audibility in the same frequency range, as shown by Motlagh Zadeh et al. (2019) and Trine and Monson (2020), who found that improved EHF audibility enhances SiN perception. Mishra et al. (2022a) reported that children with reduced sensitivity at EHF performed worse on digits-in-noise tasks compared to their peers with normal EHF hearing. However, these and other studies (Mishra et al., 2022a, 2022b; Motlagh Zadeh et al., 2019) also uncovered a secondary effect of EHFHL on SiN perception, with reduced sensitivity of the same individuals in the SF range. Motlagh Zadeh et al. (2019) and Mishra et al. (2022b) found that individuals with EHFHL had an average SF threshold approximately 4 dB poorer than those with normal EHF thresholds. These findings suggest that EHFHL might be an early marker of more distributed cochlear damage, supporting the hypothesis that OHCs in the EHF region are more vulnerable than those in the SF region (Liberman & Dodds, 1984; Le Prell et al., 2011).

Despite these findings, some studies found no significant relationship between EHF thresholds and SiN performance. Guest et al. (2018) observed small, insignificant mean differences (ranging from 3.1 dB at 9 kHz to 5.6 dB at 14 kHz) in EHF thresholds between listeners with SiN impairment and controls. Similarly, Couth et al. (2020), Badri et al. (2011), and Liberman et al. (2016) did not find significant correlations between EHF thresholds and SiN performance. These differing findings highlight the need for a targeted research approach that considers the complex role of EHF hearing in SiN perception.

### Cochlear Synaptopathy and Efferent Effects on Speech Perception in Noise

Recent evidence has implicated auditory pathways that evoke the middle ear muscle reflex (MEMR) in studies of cochlear synaptopathy due to noise, hypothesized to underlie SiN problems in humans. In rodents, a shallower MEMR input-output (growth) function is associated with synaptopathy after noise exposure (Valero et al., 2016, 2018). In humans with noise-induced tinnitus but normal hearing thresholds, a shallower MEMR growth function was also reported (Wojtczak et al., 2017). However, in humans with CF who were treated with AGs, *increased* growth of the MEMR was found while MEMR thresholds were not affected (Westman et al., 2021), suggesting altered efferent input. Considered together, there is emerging evidence for reduced MEMR growth in noise exposure, and increased MEMR growth in AG exposure. Despite hypotheses linking cochlear synaptopathy to SiN difficulties in noise and AG exposure, SiN was not tested in any of these studies. Thus, an important gap in prior research is the extent to which MEMR is related to SiN difficulty in AG ototoxicity.

#### Aim of the present study

We aimed to investigate the association between hearing damage at both SF and EHF with impaired SiN perception in children and young adults with CF who have undergone intravenous AGs treatment. We <u>hypothesized</u> that SiN perception difficulties in individuals with CF are linked to combined cochlear and synaptic or neural damage in both the SF and EHF range. We proposed three potential contributors to assess this hypothesis: 1) a primary effect of reduced EHF sensitivity, measured by pure-tone audiometry (PTA) and transient-evoked otoacoustic emissions (TEOAEs); 2) a secondary effect of subclinical damage in the SF range, measured by PTA and TEOAEs; and 3) additional neural effects, measured by MEMR threshold (afferent) and growth functions (efferent). These three contributors are not mutually exclusive; each underscores different aspects of how EHF impairment could be linked to functional hearing, suggesting that a comprehensive approach to auditory testing could be crucial for diagnosing AG-related ototoxicity.

## Methods

### Participants

The study included 101 ears from participants with CF treated with intravenous AGs (tobramycin or amikacin, with or without vancomycin) and 84 ears from age- and sex-matched non-CF (Control) participants with no history of AGs exposure.

Participants ranged in age from 7 to 21 years (M = 15.32, SD = 3.13; 58% female). Individuals with CF were recruited from the CF inpatient unit at Cincinnati Children’s Hospital Medical Center (CCHMC), while control participants were recruited from the same hospital through internal staff emails and outpatient study flyers. Additional eligibility criteria for both the CF and control groups included the ability to complete a conventional behavioral hearing test and being a native English speaker. Controls were not excluded based on hearing status to ensure representation of the general population’s hearing levels. Subjects with middle ear disorders/conductive hearing loss were excluded. Informed patient consent (for ages 18 years old or older) and parental consent and child assent (for ages 11 -17 years old) were obtained for all participants before enrollment. All participants were reimbursed for their participation.

### Procedures

Comprehensive assessments were performed on both test groups to evaluate their hearing threshold, cochlear function, and speech perception.

#### Audiometric assessment (Pure-Tone Hearing Thresholds)

Air conduction (AC) hearing thresholds were obtained using an Equinox Audiometer (Interacoustics, Inc.) and Sennheiser HDA-300 circumaural headphones. Frequencies tested included octave SF ranging from 0.25 to 8 kHz and EHF measured at 10, 12.5, 14, and 16 kHz. Calibration was completed according to ISO 389.8 (International Organization for Standardization, 2004) for SF and ISO 389–1 (International Organization for Standardization, 2017) for EHF. If AC thresholds were ≥ 20 dB HL at 0.25, 0.5, 1, 2, or 4 kHz, bone conduction thresholds were obtained utilizing a RadioEar B–71 bone oscillator placed at the mastoid (Radio-Ear Corp.) with masking applied to the contralateral ear. Hearing loss was defined as audiometric thresholds > 20 dB HL in either ear at any frequency. For statistical analysis, hearing threshold data for each ear were categorized into three measures: Low-Frequency Pure-Tone Average (LF-PTA_(0.25, 0.5, 1 kHz)_), High-Frequency PTA (HF-PTA_(2, 4, 8 kHz)_), and EHF-PTA_(10, 12.5, 14, 16 kHz)_.

#### Middle Ear Measure (Tympanometry)

Traditional 226-Hz tympanometry was conducted to assess middle ear admittance using the Interacoustics Inc. Titan. Abnormality in conductive hearing was determined based on two or more measures (admittance, tympanometric peak pressure, and tympanometric width) falling outside normative ranges (admittance between 0.3 and 1.5 mmho, peak pressure between −100 and +30 daPa, and tympanometric width between 30 and 105 daPa).

#### Middle Ear Muscle Reflex

MEMRs were assessed using the wideband absorbance method, implemented via custom MATLAB software as outlined by Keefe et al. (2017). The probe assembly included a high-bandwidth receiver for delivering wideband clicks as the probe stimulus, and a second receiver with the same bandwidth to accommodate higher stimulus levels. Stimulus level calibration was achieved using a 2-cc coupler. Broadband noise (BBN, 0.2 to 8 kHz) was presented to the probe ear in the ipsilateral mode and the opposite ear in the contralateral mode, while wideband clicks were delivered to the probe ear to monitor absorbance changes. The ear canal pressure was set to the average peak tympanometric pressure obtained from both down-swept and up-swept wideband tympanograms. Both contralateral and ipsilateral MEMR testing incorporated response averaging, artifact rejection, and signal processing to evaluate reflex threshold, onset latency, and amplitude growth (Keefe et al., 2017). MEMR responses to broadband noise (BBN) elicitors were measured from 50 to 90 dB SPL in 5 dB increments, corresponding to 10 stimulus levels (L1–L10, with L1 representing the lowest level). At each level, MEMR amplitude was quantified as the cumulative change in absorbed power level (dB), averaged across lower frequencies (0.2–2.4 kHz) where reflex activity is most prominent (Hunter et al., 2023). MEMR growth was defined as the slope of the reflex amplitude function across elicitor levels, representing the rate of change in MEMR amplitude (Δ absorbance in dB) per increase in stimulus level (dB SPL). This approach captures the change in reflex amplitude across the tested intensity range (Vasudevamurthy et al., 2023). For statistical analyses, MEMR threshold and MEMR growth slope were used as predictors in the models.

#### The Bamford–Kowal–Bench Speech-in-Noise (BKB-SIN) Test

To assess speech perception in noise, the Bamford-Kowal-Bench Speech-in-Noise (BKB-SIN) Test (Etymotic Research, 2005) was administered at 50 dB HL in a monaural setup, with one list pair presented to each ear. The same circumaural earphones as for audiometry were used (Sennheiser HDA 300). This adaptive test adjusts to a range of signal-to-noise ratios (SNR) from +21 to −6 dB, reflecting typical classroom and everyday noise conditions. The BKB-SIN sentences, recorded at a 44.1-kHz sampling rate, have significant signal energy up to 12 kHz. Participants were asked to repeat each sentence and guess if it is uncertain. The number of correctly repeated keywords was used to determine the SNR necessary for the participant to understand 50% of the sentences (SNR-50). Signal-to-noise ratio loss (SNR-Loss) was calculated by subtracting age-matched normative SNR-50 values from the individual’s SNR-50. This calculation shows how much higher the SNR must be for the participant to achieve performance comparable to their age-matched peers (Etymotic Research, 2005). In this study, SNR-Loss was utilized as the primary outcome variable in each ear.

#### Transient-evoked otoacoustic emission

TEOAEs were utilized to evaluate OHC function in the cochlea, focusing on reflection-based emissions. A specialized system that uses chirp stimuli (ranging from low to high frequencies) combined with double-evoked techniques was employed to allow broader frequency recording from 0.7 to 14.7 kHz than is possible using clinical TEOAE systems (Keefe et al. 2019). The double-evoked technique eliminates stimulus artifacts, allowing for accurate recording at higher frequencies. Additionally, chirp stimuli, which have a longer duration compared to click stimuli, reduce distortion at elevated intensity levels (Keefe et al., 2019). Two chirp stimuli were utilized: the first addressed the SF (0.5-8 kHz, at 78 dB peSPL) and the second focused on EHFs (8-14.7 kHz, at 82 dB peSPL). Both chirp stimuli were presented at a sweep frequency rate of 188 Hz/ms. TEOAE responses were recorded using an Etymotic ER10B+ microphone, coupled with ER2 sound sources and a sound card operating at a 48 kHz sample rate (RME Babyface), under the control of a custom MATLAB program. For analysis, TEOAE-SNR data for each ear were categorized into three average measures: LF-SNR_(0.75, 1 kHz)_, HF-SNR_(2, 4, 8 kHz)_, and EHF-SNR_(10, 12.7, 14.2kHz)_.

### Statistical Analysis

Descriptive statistics were used to summarize demographic variable measurements. Histograms and scatter plots were utilized to examine data distribution and relationships among variables. Spearman correlation coefficients were calculated to explore associations among variables. To study the effects of demographic and exploratory variables on the outcome measure (BKB-SNR Loss), while accounting for repeated measures from both ears, mixed-effects models were performed. An initial unadjusted univariate analysis was conducted for each demographic or explanatory variable. Predictors with at least marginal significance (*p* < 0.1) were included in subsequent adjusted analyses. Multicollinearity among the predictor variables was assessed using correlation coefficients and the Variance Inflation Factor. Data analysis was performed using SAS statistical software, version 9.4 (SAS Institute, Cary, N.C.), with a significance level set at 0.05.

## Results

### Overall Group Differences in EHF Hearing and SiN Performance

Of the 185 ears analyzed, 34 ears (18.4%) showed EHFHL, while 151 ears (81.6%) had normal EHF hearing (NEHF). EHF hearing thresholds were significantly higher in the CF group than in controls (β = −9.98, t(183) = −4.73, *p* < 0.001; Figure 1). Additionally, a chi-square test revealed a significant association between participant group (CF vs. control) and EHF hearing status, χ²(1, N = 185) = 10.35, *p* = .001, indicating that individuals with CF (27%) were more likely to exhibit EHFHL than controls (6%).

**Figure 1.**
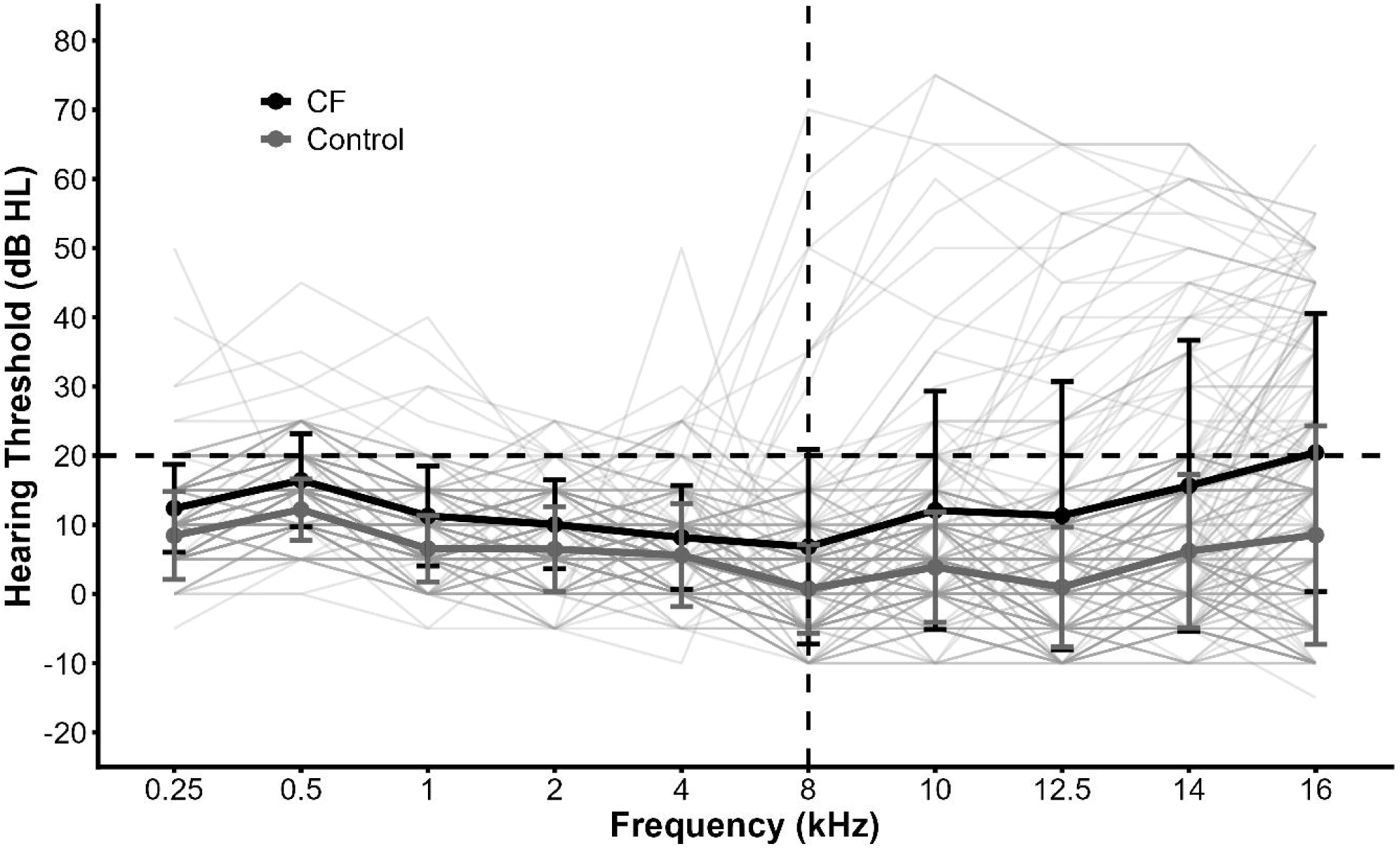
Pure-tone audiometric thresholds across frequency for ears from participants with cystic fibrosis (CF) and controls. Thin gray lines represent individual ears, while colored lines and error bars indicate group means ±1 SD. The vertical dashed line denotes the 8 kHz boundary between standard and extended high frequencies.

Participants with CF exhibited greater BKB-SNR loss than controls, and group (CF vs. control) was a significant predictor of BKB-SNR loss in a linear mixed-effects model accounting for repeated measures from both ears (β = 1.48, *p* < .0001; Figure 2).

**Figure 2:**
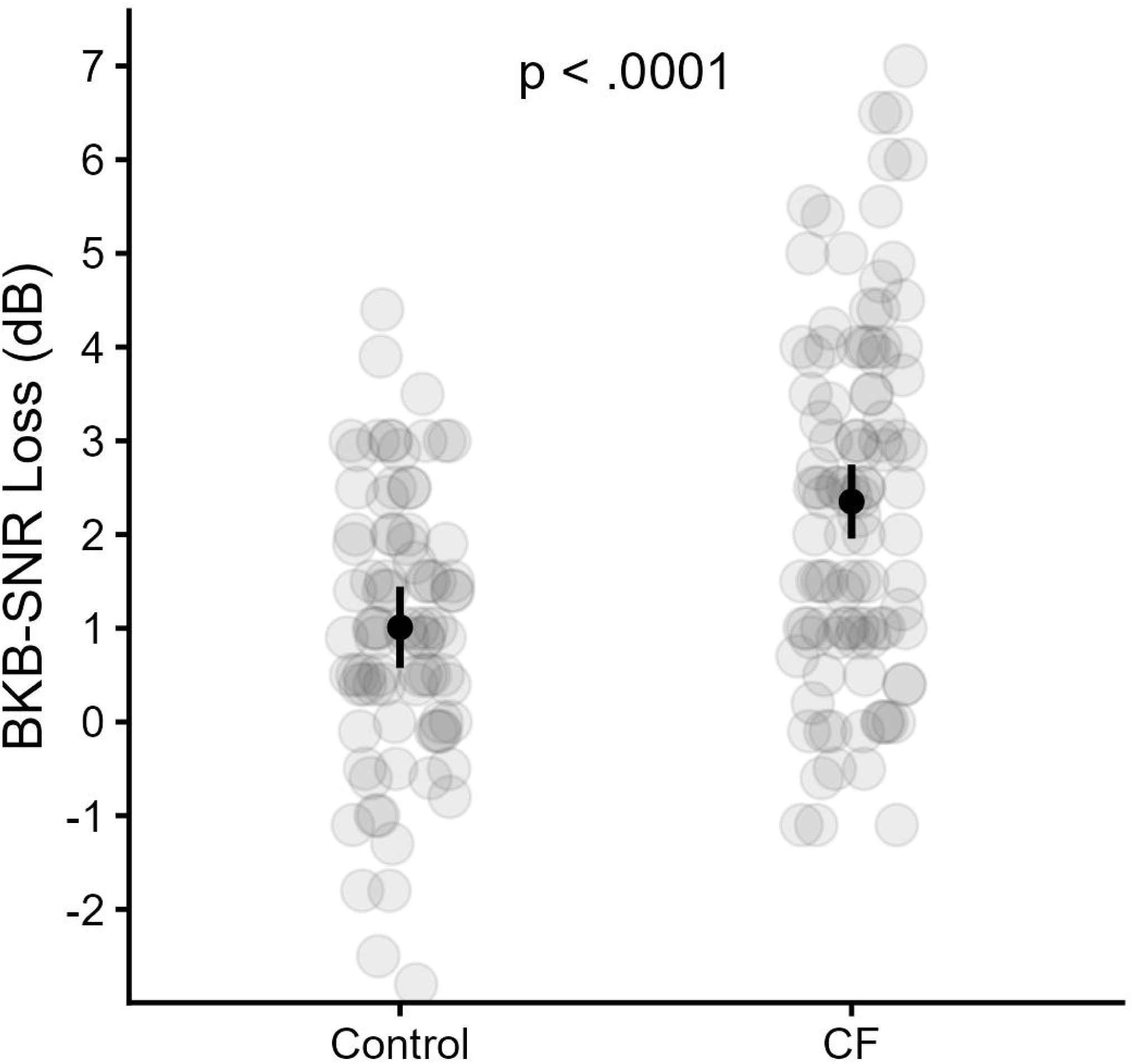
BKB-SNR loss for the cystic fibrosis (CF) and control groups. Gray circles represent individual ears. Black points indicate group means with 95% confidence intervals.

To further characterize cochlear function associated with EHFHL, correlations were examined between EHF hearing status and audiometric and TEOAE measures across ears from both CF and control groups. Spearman correlations showed that ears with EHFHL had significantly poorer LF-PTA (Figure 3a; ρ = 0.41, *p* < .0001) and HF-PTA (Figure 3b; ρ = 0.46, *p* < .0001). As expected, ears with EHFHL also demonstrated reduced TEOAE SNR at EHF (ρ = - 0.19, *p* = .01), LF (ρ = - 0.18, *p* = .01), and HF (ρ = - 0.28, *p* < .0001) relative to ears with NEHF (Figures 4a-4c). No significant associations (all *p* > .05) were observed between EHF hearing status and age or sex.

**Figure 3:**
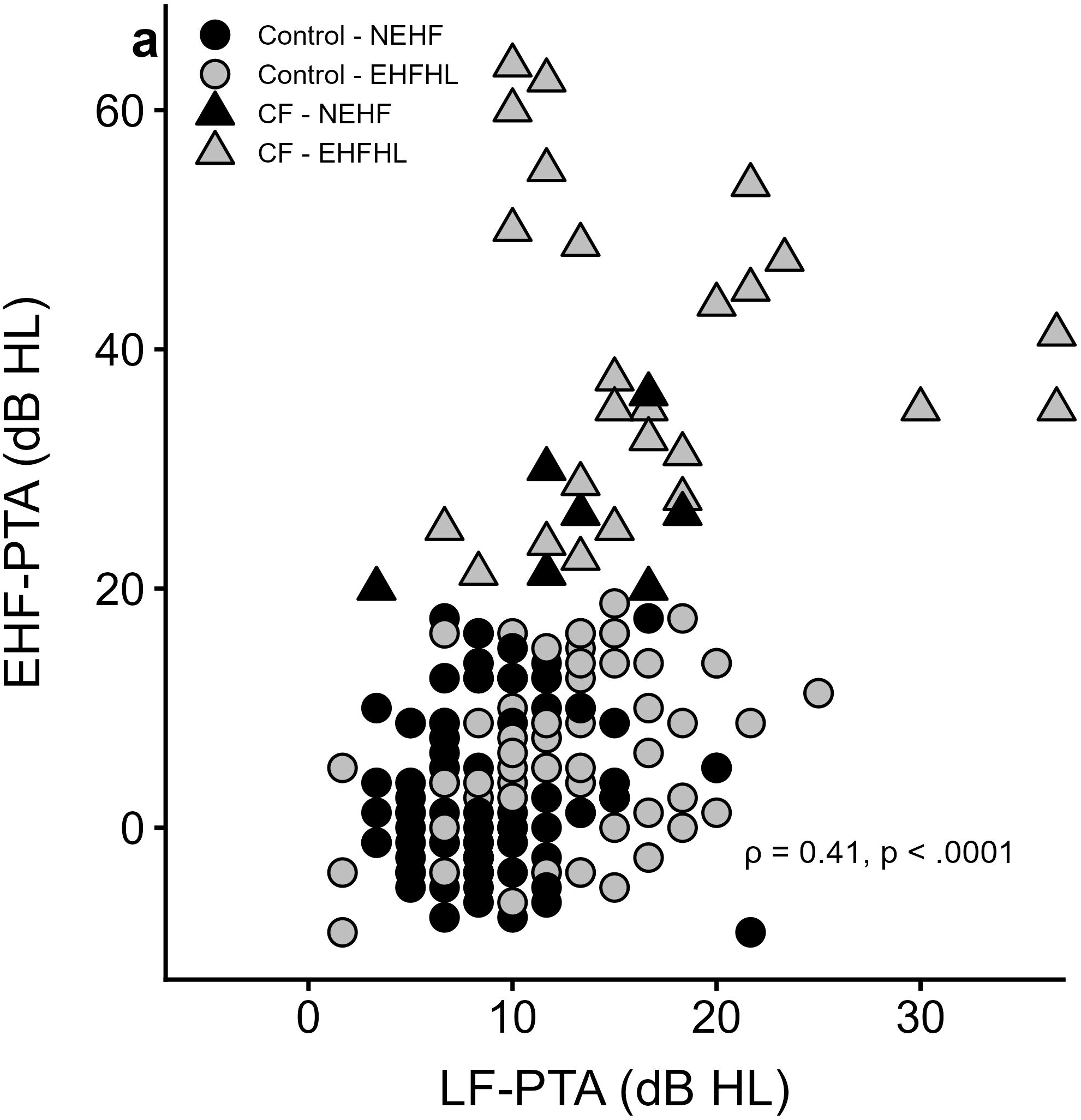
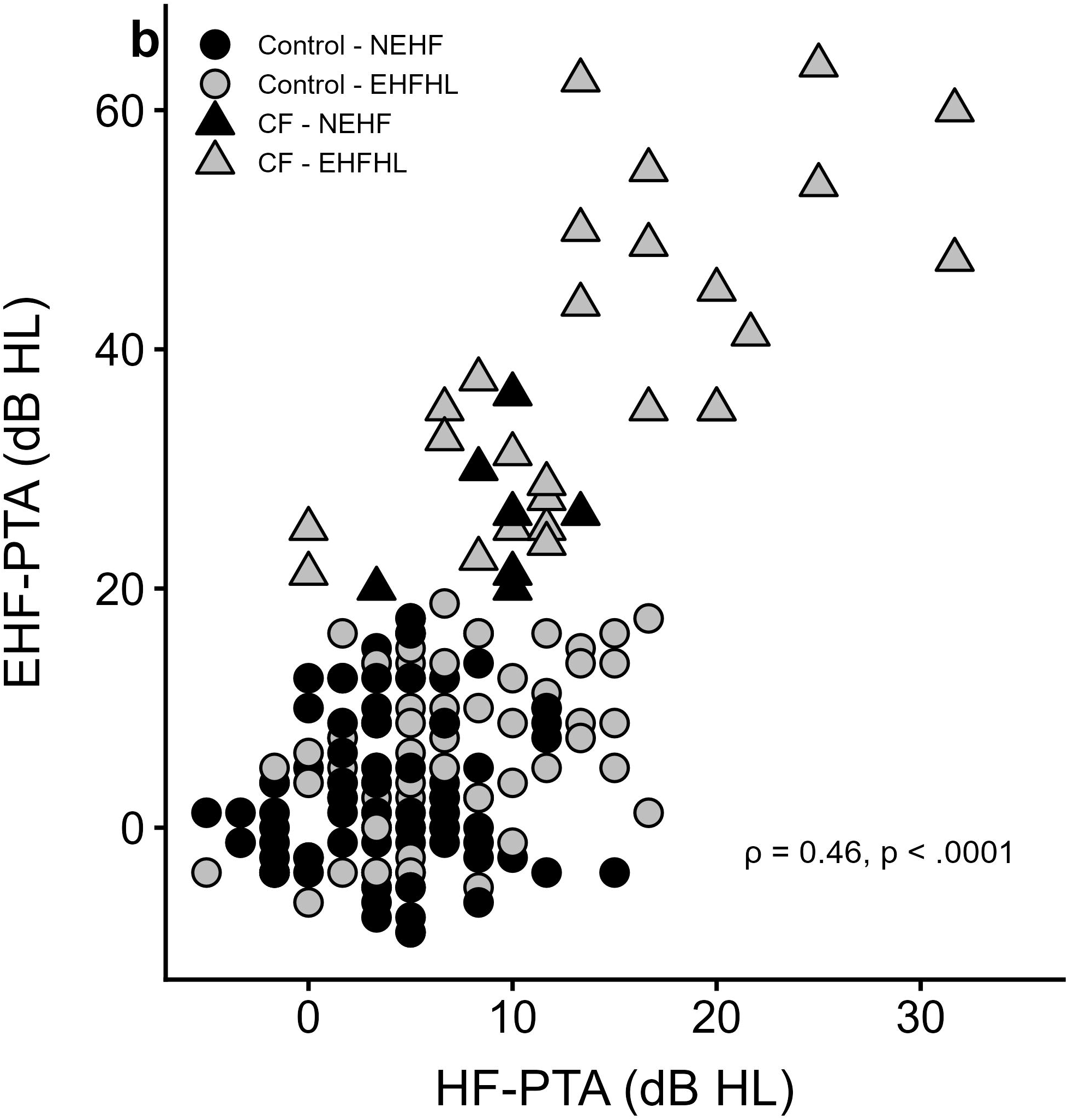
Association between extended high-frequency pure-tone average (EHF-PTA) and **(a)** low-frequency pure-tone average (LF-PTA) and **(b)** high-frequency pure-tone average (HF-PTA). Each point represents an ear. Point shading denotes EHF hearing status (normal EHF [NEHF] vs. extended high-frequency hearing loss [EHFHL]), and symbol shape denotes participant group (CF vs. control). Spearman correlations showed significant positive relationships in both panels.

**Figure 4:**
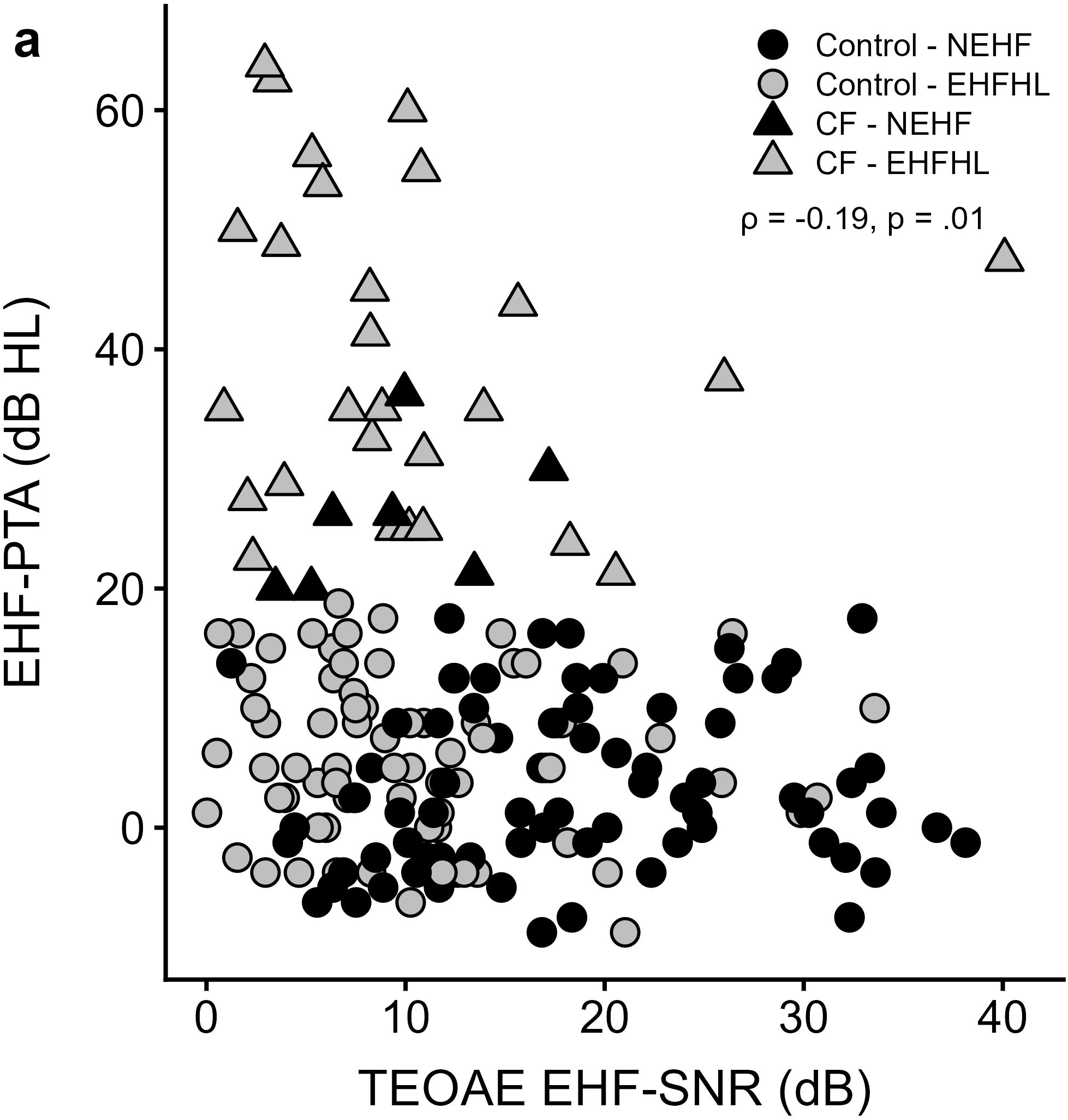
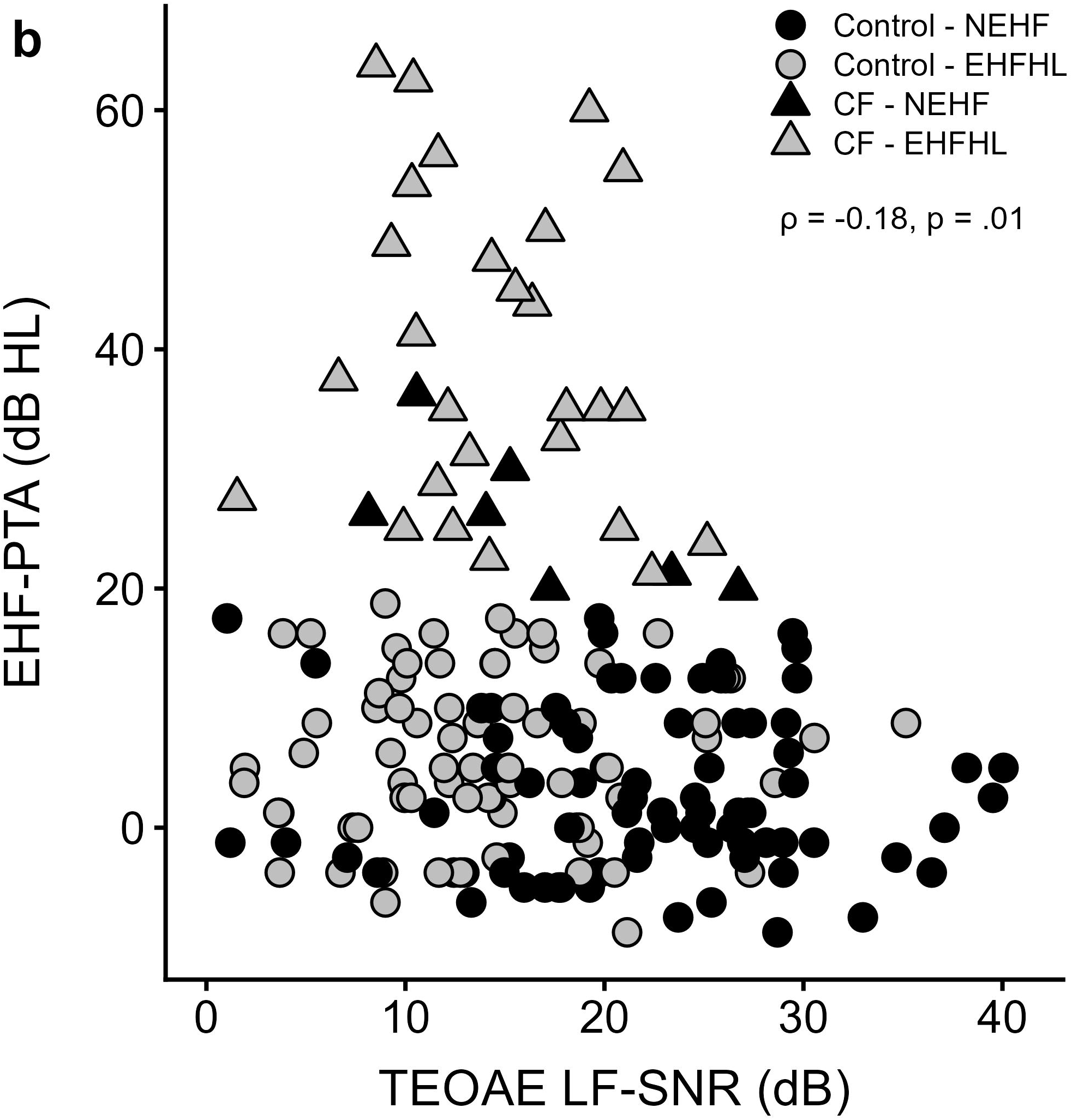
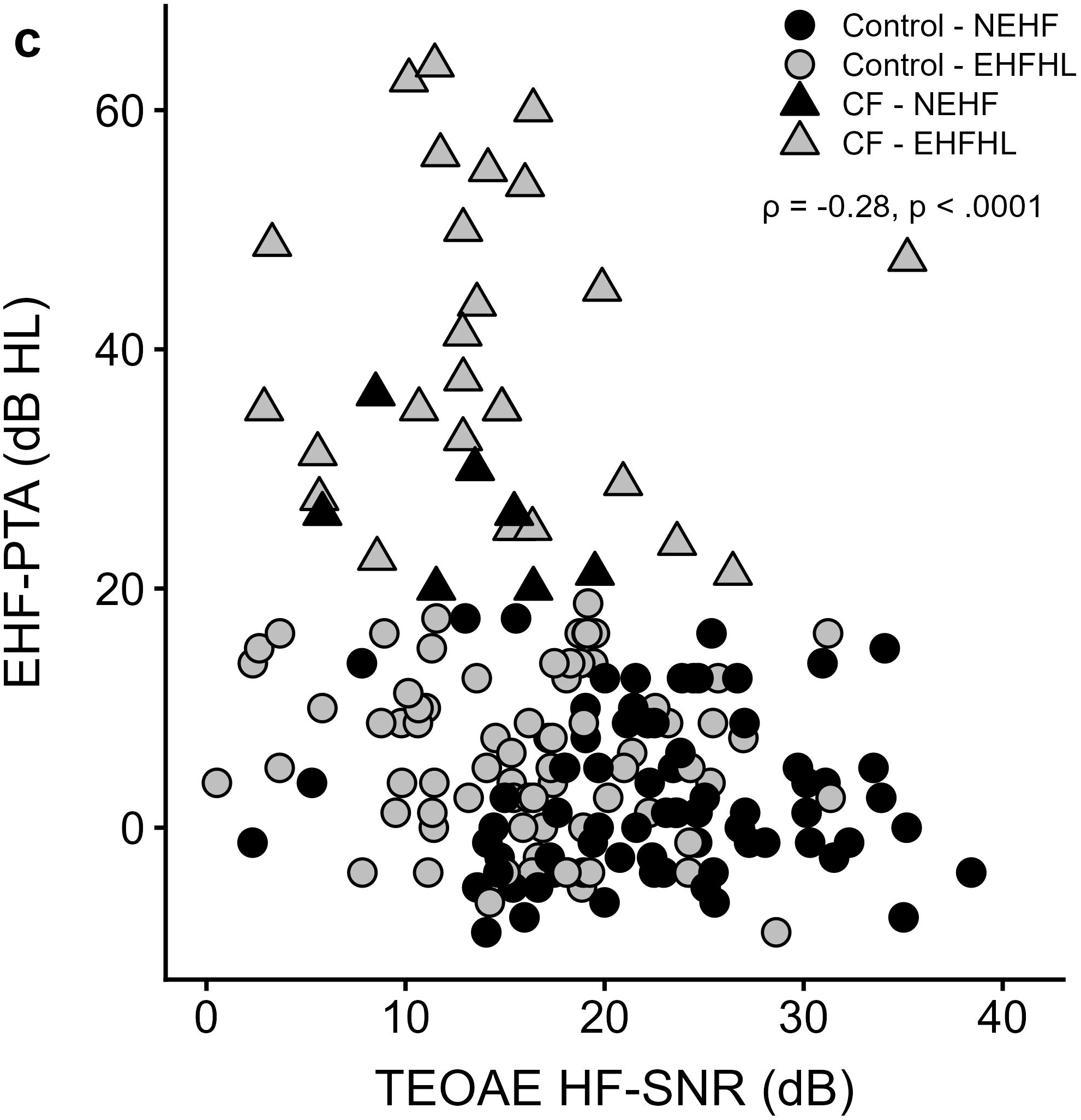
Association between extended high-frequency pure-tone average (EHF-PTA) and TEOAE signal-to-noise ratio (SNR) at **(a)** EHF; **(b)** low-frequency (LF); and **(c)** high-frequency (HF). Point shading denotes EHF hearing status (normal EHF [NEHF] vs. EHF hearing loss [EHFHL]), and symbol shape denotes participant group (CF vs. control). Spearman correlations indicated significant negative relationships in all panels.

The following sections present results for each of the three hypothesized contributors to SiN perception:

**1) Primary effect of EHFHL on SiN perception**

Contrary to our primary hypothesis, in both univariate and multivariate models, EHF hearing status did not significantly influence BKB-SNR Loss (F(1,10) = 0.24, *p* = .63).

Additionally, EHF-PTA (F(1,89) = 2.04, *p* = .15) and TEOAE EHF-SNR (F(1,89) = 1.43, *p* = .23) were not significantly associated with BKB-SNR Loss. Participant group (CF vs. control) did not significantly interact with these predictors (all *p* > .05).

**2) Secondary effect of EHFHL on SiN perception mediated by SF hearing**

SF hearing measures were significantly associated with SiN performance. In univariate analyses, LF-PTA (F(1,89) = 9.17, *p* = .003), HF-PTA (F(1,89) = 14.02, *p* = .0003), and TEOAE LF-SNR (F(1,89) = 5.48, *p* = .02) were significant predictors of BKB-SNR Loss, suggesting that elevated thresholds and reduced OHC responses in the SF range were linked to poorer SiN performance. Multivariate mixed-effects models confirmed that LF-PTA (*p* = .01) and HF-PTA (*p* = .008) remained significant predictors, with each 1-unit increase associated with 0.07 and 0.11 increases in BKB-SNR Loss, respectively (Figures 5a and 5b). Moreover, TEOAE LF-SNR (*p* = .05) and HF-SNR (*p* = .008) approached significance, with higher SNR values (better OHC function) predicting lower BKB-SNR Loss (0.043 and 0.065 per unit, respectively; Figures 6a and 6b).

**Figure 5:**
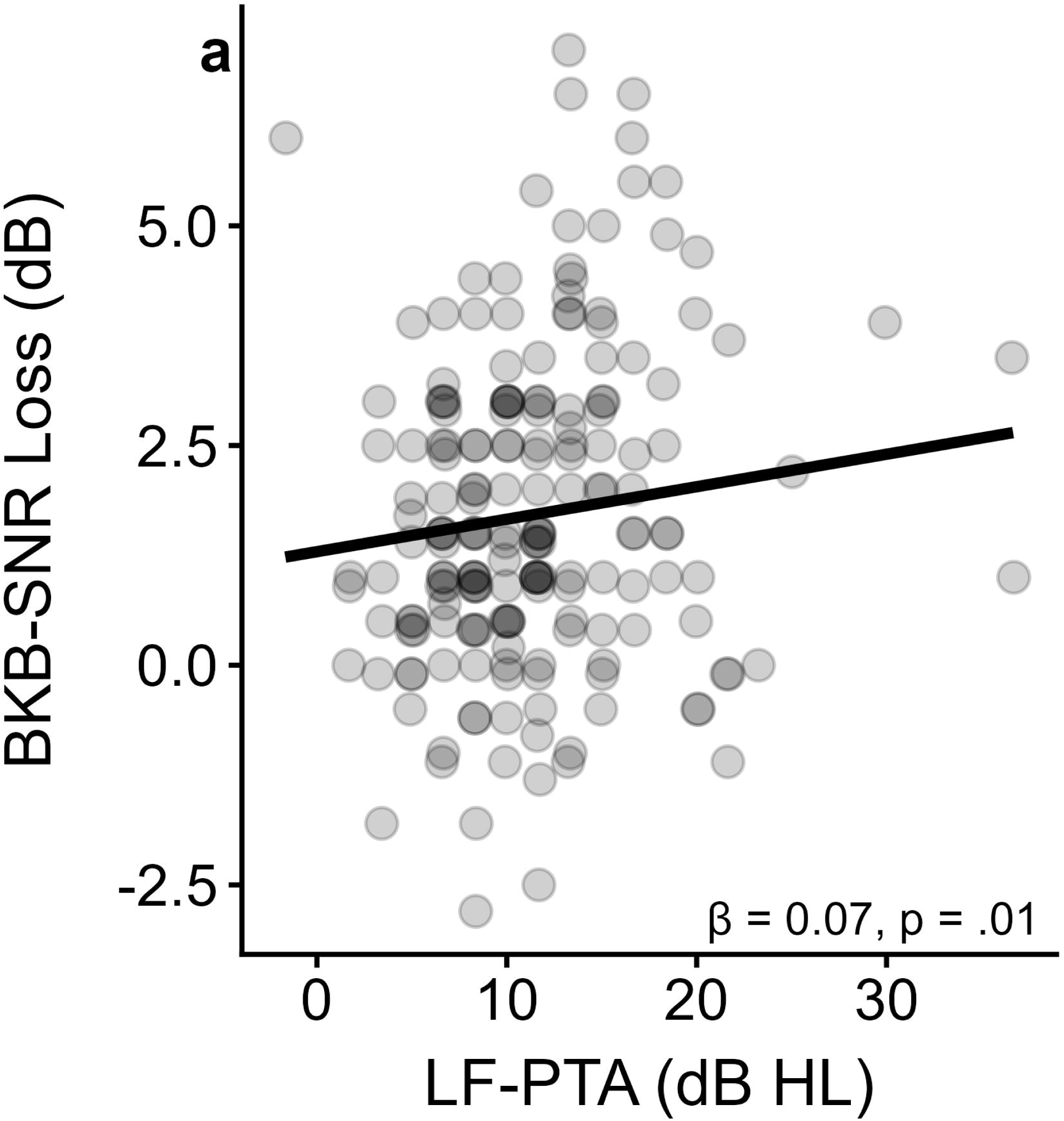
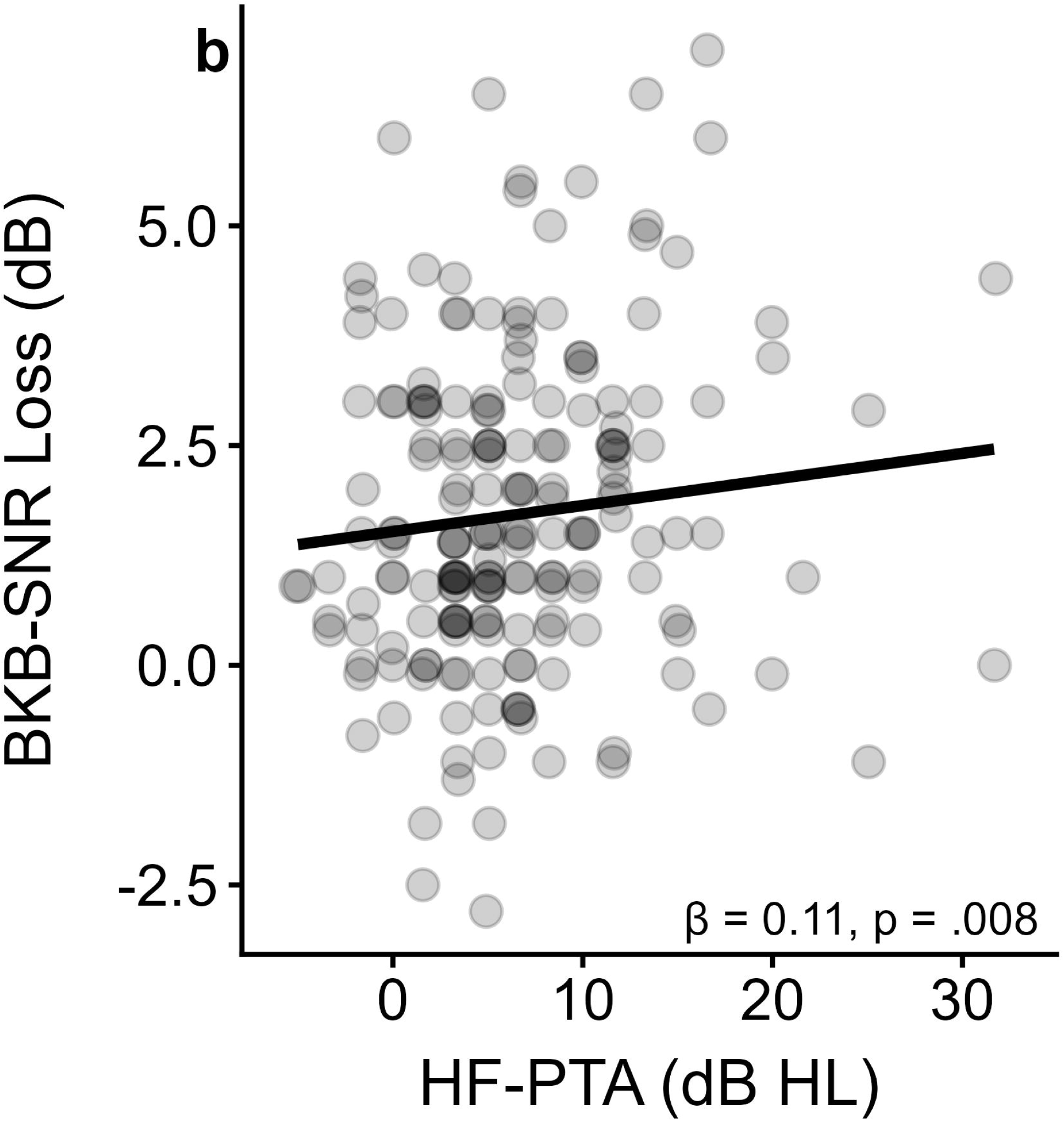
Associations between pure-tone thresholds and speech-in-noise performance. Scatter plots show individual BKB-SNR Loss values (faded points). Solid lines represent marginal predictions from the multivariate mixed-effects model.

**Figure 6:**
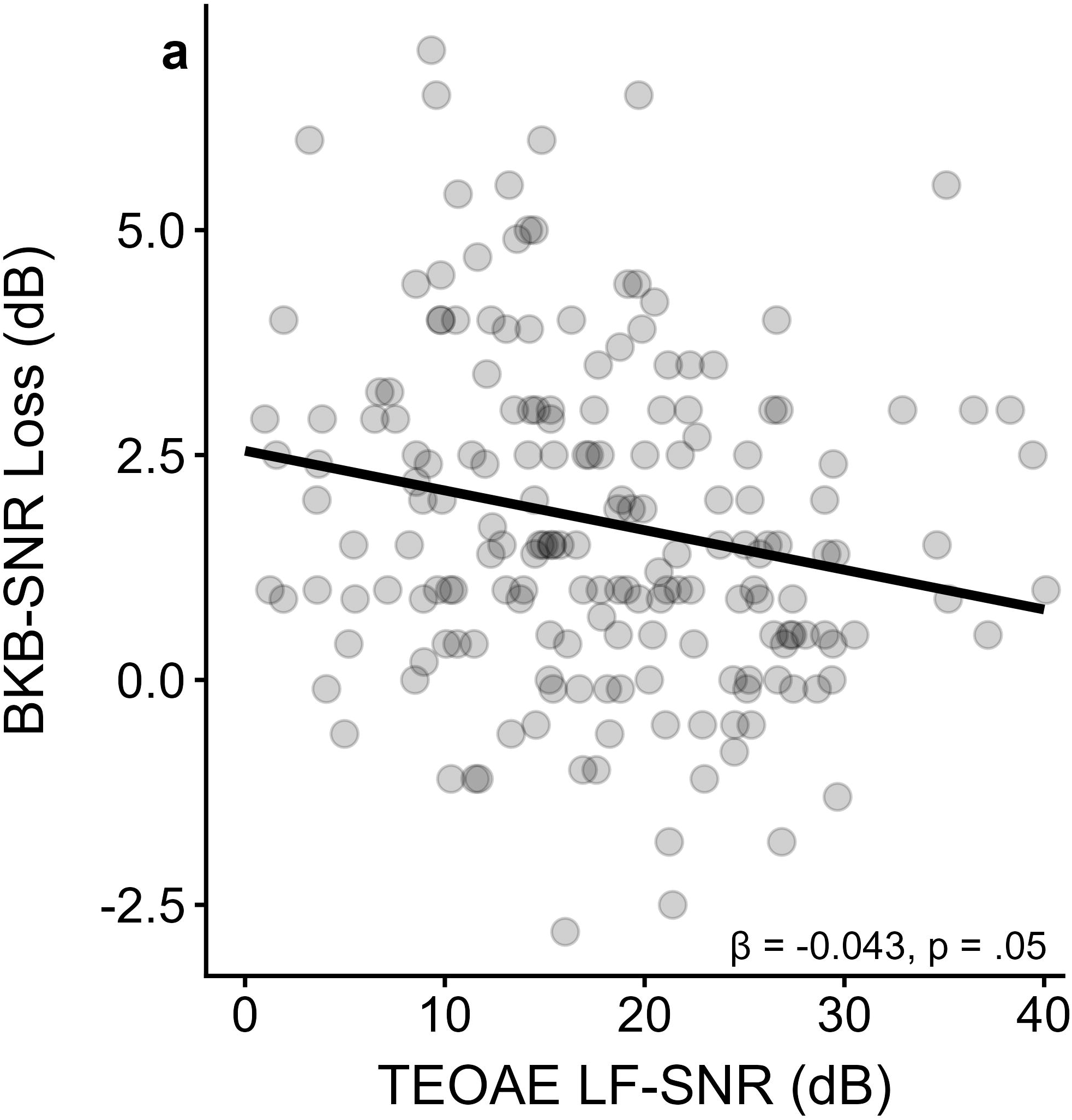
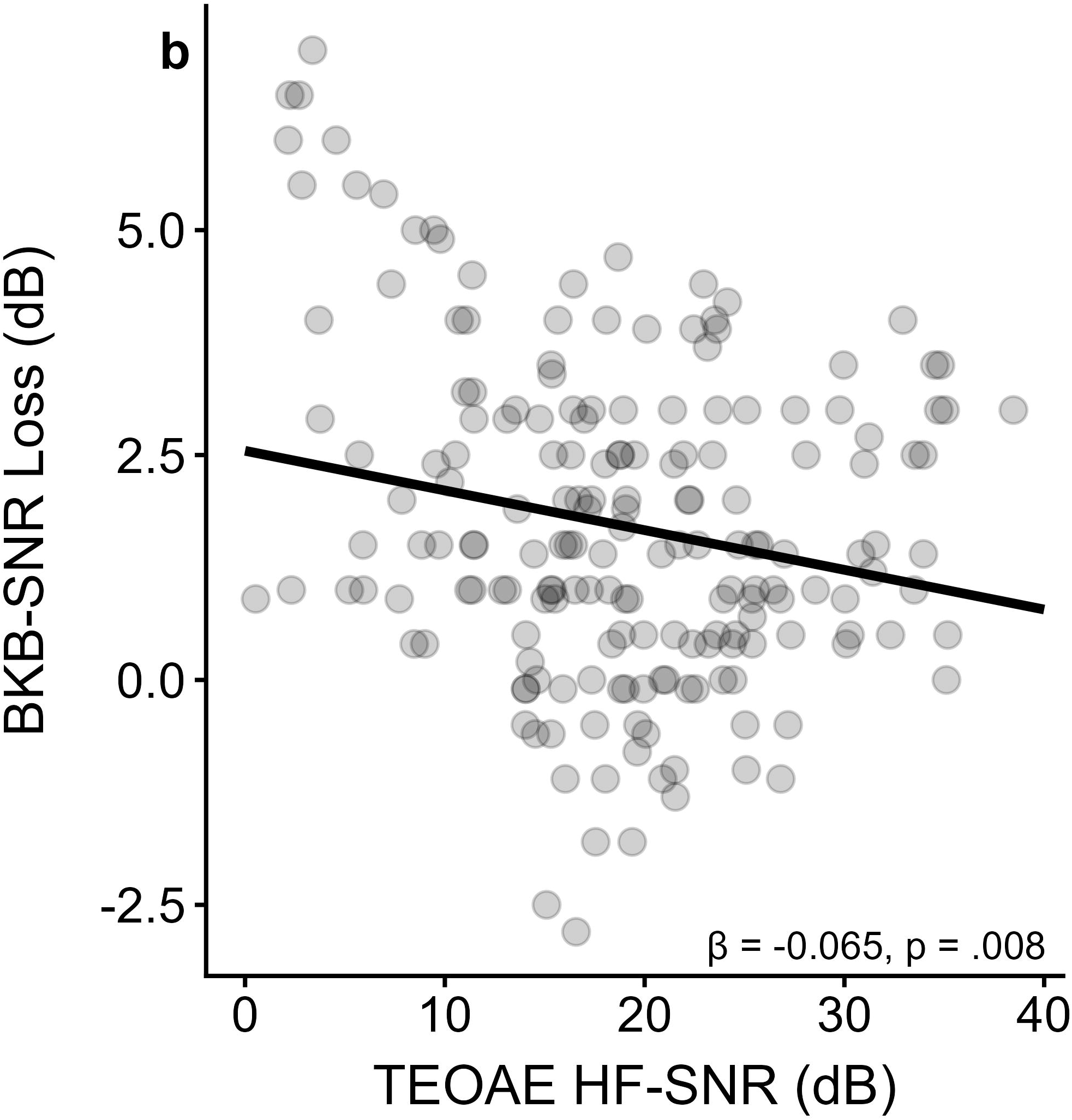
Associations between TEOAE signal-to-noise ratio (SNR) and speech-in-noise performance. Scatter plots show individual BKB-SNR Loss values (faded points). Solid lines represent marginal predictions from the multivariate mixed-effects model.

These relationships did not differ by participant group (CF vs. control; all interaction *p* > .05).

**3) Synaptic (afferent) or efferent contributions reflected by MEMRs**

EHF-PTA and TEOAE EHF-SNR measures were not significantly associated with MEMR measures (all *p* > .05), suggesting that EHF hearing loss alone did not account for MEMR variability. In contrast, MEMR growth functions were significantly associated with BKB-SNR loss, with significant interactions with participant group. For contralateral MEMR growth, the overall model was significant (F(1,76) = 4.8, *p* = .01), driven primarily by the interaction with group (F(1,76) = 5.3, *p* = .02), with the CF group exhibiting a slope 0.34 units steeper than controls (Figure 7a). Similarly, ipsilateral MEMR growth was significant overall (F(1,80) = 6.75, *p* = .001), largely reflecting the interaction with group (F(1,80) = 7.9, *p* = .006), with CF participants showing a slope 0.36 units steeper than controls (Figure 7b). These findings indicate a greater increase in MEMR amplitude with increasing stimulus level in the CF group. MEMR threshold measures were not significantly associated with BKB-SNR Loss or any other predictors in the models (all *p* > .05).

**Figure 7:**
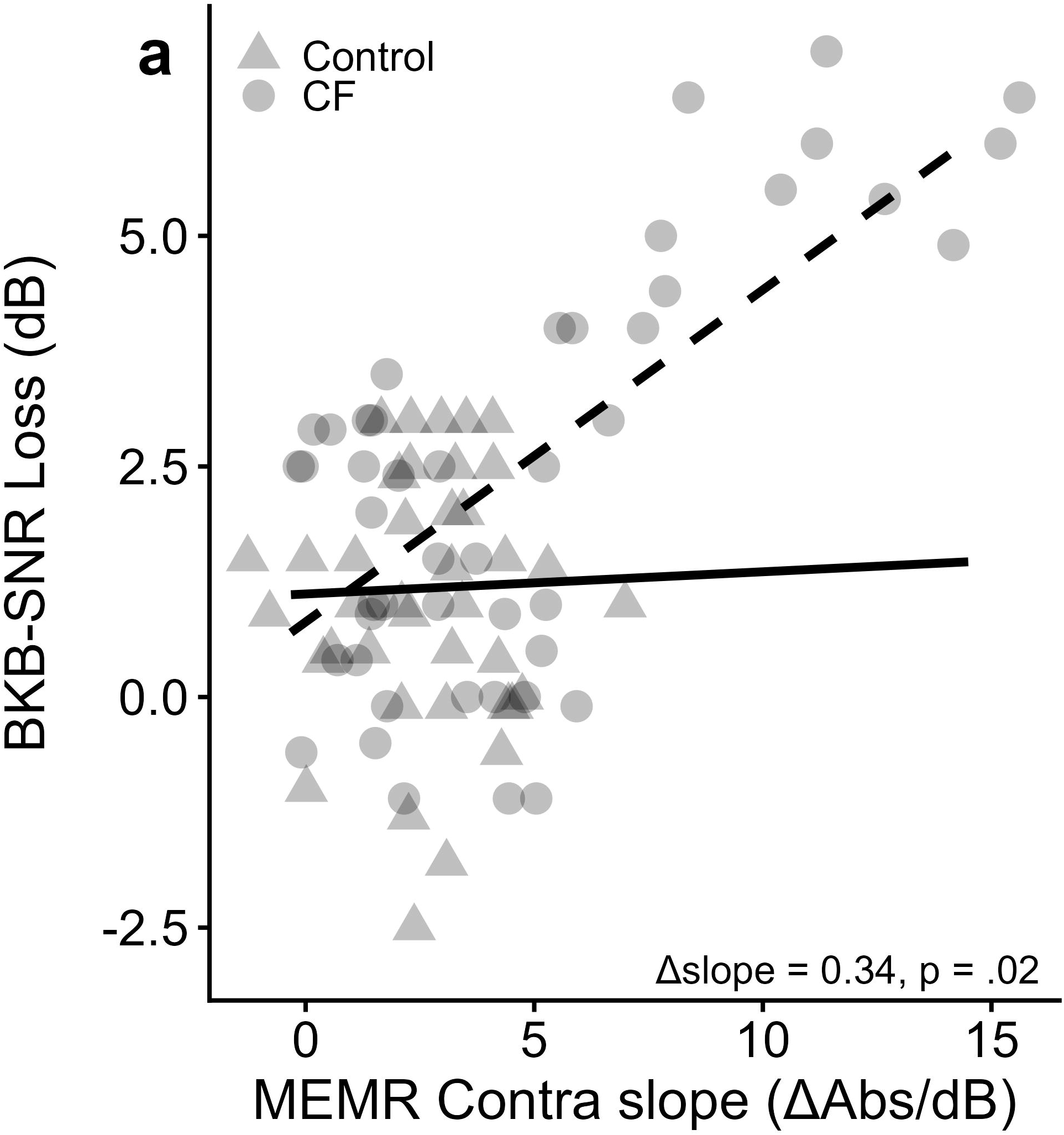
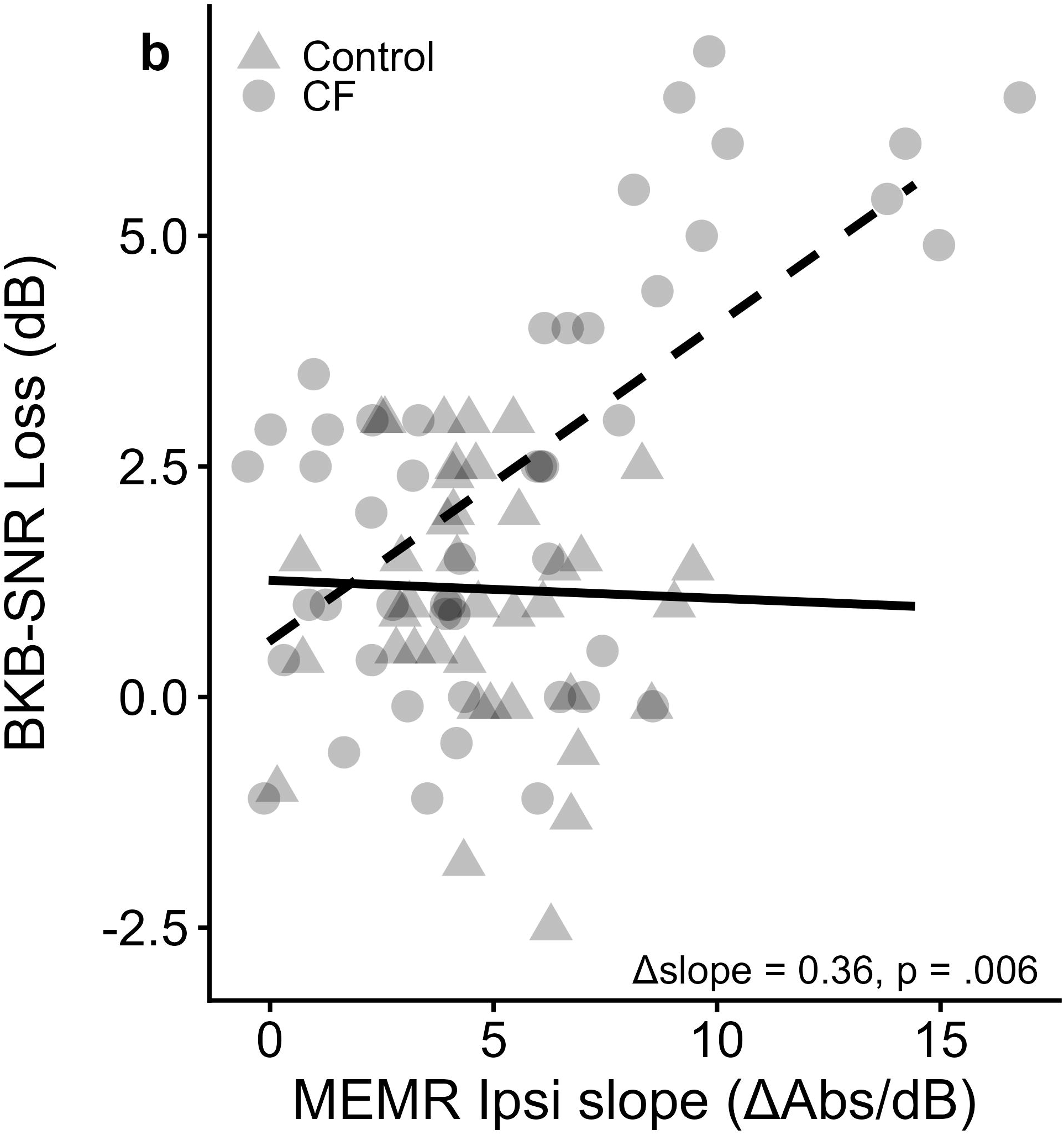
Associations between MEMR growth slope and BKB-SNR loss for **(a)** contralateral and **(b)** ipsilateral conditions. Solid lines indicate model-predicted trends for the Control group, and dashed lines indicate model-predicted trends for the CF group. Δslope values reflect the difference in regression slopes between CF and Control groups derived from linear mixed-effects models. ΔAbs/dB = change in reflex absorbance per dB increase in elicitor level.

## Discussion

The present study evaluated sensory and neural contributions to SiN perception in children and young adults with CF exposed to intravenous AGs. Although EHFHL is well documented as an early manifestation of AG ototoxicity (Fausti et al., 1994; Konrad-Martin et al., 2005; Hinojosa et al., 2001), the functional consequences of this early damage for everyday communication remain unclear. Histopathologic and animal studies show that aminoglycoside toxicity typically begins in the basal cochlea, where outer hair cells encoding extended high frequencies are particularly vulnerable (Forge & Schacht, 2000; Xie et al., 2011). However, these same studies also demonstrate that AG exposure can affect inner hair cell synapses and spiral ganglion neurons, producing neural degeneration that extends beyond the region of initial hair cell loss (Hinojosa et al., 2001; Sergeyenko et al., 2013). Such neural injury may occur even when audiometric thresholds remain relatively preserved. In this context, our findings indicate that EHF hearing status alone does not explain SiN difficulties in this population.

Instead, subtle deficits in SF hearing and enhanced MEMR growth dynamics were the strongest predictors of SiN performance, consistent with a broader pattern of cochlear and neural involvement. Together, these results suggest that the auditory phenotype associated with AG exposure in CF reflects interacting sensory and neural mechanisms rather than isolated basal cochlear damage.

### Primary effect of EHFHL on SiN

Although EHFHL and SiN loss were both more prevalent in the CF cohort, EHF thresholds and EHF TEOAEs were not directly associated with SiN performance. This pattern aligns with several studies reporting minimal or inconsistent contributions of EHF sensitivity to sentence-level SiN recognition (Guest et al., 2018; Couth et al., 2020; Liberman et al., 2016). The sentence task in competing noise with monaural presentation was designed to tap functional communication demands in relation to hearing sensitivity in each ear individually. However, methodological and perceptual factors may have limited the observable impact of EHF cues in the present paradigm.

First, although BKB-SIN stimuli contain measurable spectral energy extending to approximately 10-12 kHz, most of the intelligibility-relevant phonetic information, at least for adults, exists below 8 kHz (Monson et al., 2014). The effective speech energy within the 8-12 kHz band may therefore have been insufficient for EHF sensitivity to influence performance. Second, sentence-level tasks carry substantial linguistic redundancy and provide listeners opportunities to compensate for reduced high-frequency detail using top-down contextual cues (Leibold et al., 2019; Buus et al., 1986). EHF contributions may be more apparent in tasks with minimal linguistic support, such as digits-in-noise or phoneme identification (e.g., Mishra et al., 2022a; Vitela et al., 2015). Third, the monaural headphone presentation eliminated spatial hearing cues that enhance speech-noise segregation and may increase the perceptual weight of high-frequency information. Finally, the BKB-SIN test uses relatively low-energetic masking noise, reducing spectral competition at high frequencies and thereby limiting the need for fine spectral resolution in the EHF range. Collectively, these tasks and stimulus characteristics likely reduced the perceptual demands on EHF hearing and help explain the absence of a direct EHF-SiN association in this study.

### Secondary SF Effects of EHFHL on SiN

Although EHF sensitivity did not directly predict SiN, ears with EHFHL also exhibited poorer SF thresholds and reduced SF-TEOAE amplitudes, consistent with reports that EHFHL may mark a broader cochlear vulnerability extending toward apical regions (Motlagh Zadeh et al., 2019; Blankenship et al., 2021; Jain et al., 2022; Mishra et al., 2022a and 2022b). This pattern supports the long-standing hypothesis that OHCs in the basal cochlea are most susceptible to AG-induced oxidative stress and that injury may progress apically with cumulative exposure (Kusunoki et al., 2004; Ishikawa et al., 2019). Importantly, SF hearing sensitivity emerged as the strongest predictor of SiN performance, with both LF-PTA and HF-PTA contributing significantly, and SF-TEOAE amplitudes showing additional predictive value. These findings align with extensive literature demonstrating that even mild SF threshold elevations, often within the clinically “normal” range, can degrade SiN perception due to reduced audibility and impaired spectral-temporal processing (Best et al., 2005; Hunter et al., 2020; Flaherty et al., 2021). In the context of CF, our findings extend those of Blankenship et al. (2021), who demonstrated that CF patients with AG exposure and EHFHL had poorer SiN performance, although SF thresholds remained largely normal. Because their study did not directly compare SF and EHF predictors of SiN, the present results add new evidence that SF cochlear involvement plays a primary role in functional listening outcomes. Overall, these findings suggest that EHFHL may serve as an early biomarker of cochlear vulnerability, but functional communication difficulties emerge primarily when damage extends into the SF region.

### Neural Contributions Reflected in MEMRs

MEMR growth functions were significantly associated with SiN performance, with CF ears demonstrating steeper growth slopes and stronger associations with BKB-SNR loss than controls. In contrast, MEMR thresholds were not significantly related to SiN performance, suggesting that the observed effects are unlikely to reflect differences in afferent reflex activation alone. Instead, these findings point toward enhanced efferent changes in reflex amplitude with increasing stimulus level, which are thought to reflect central gain regulation within the auditory brainstem (Liberman et al., 2016; Valero et al., 2018). Moreover, these effects occurred independently of EHF or SF sensitivity, suggesting a contribution from neural mechanisms rather than OHC dysfunction alone. Consistent with this interpretation, prior work in adults with CF demonstrated enhanced MEMR growth functions despite largely preserved audiometric thresholds, supporting the presence of altered brainstem-level auditory processing in this population (Westman et al., 2021).

Wideband MEMRs reflect the integrity of both afferent auditory nerve fibers and efferent brainstem pathways involved in gain control, acoustic scene analysis, and protection from noise (Bochat et al., 2022; Vasudevamurthy & Kumar, 2023). Altered reflex growth has been proposed as a noninvasive marker of cochlear synaptopathy or neural dysregulation (Liberman et al., 2016; Viana et al., 2015). Prior work in children with listening difficulties demonstrates similar associations between increased MEMR reflex growth function and SiN deficits (Hunter et al., 2023). Given evidence that AGs can damage spiral ganglion neurons and synapses in addition to hair cells (Sone et al., 1998; Hinojosa et al., 2001), our findings support the possibility that enhanced MEMR growth reflects altered neural gain or brainstem reflex regulation, contributing to listening challenges in CF and interacting with SF sensory loss to degrade communication outcomes.

### Limitations and Future Directions

Because the design was cross-sectional, the temporal sequence of cochlear and neural changes cannot be determined. Longitudinal studies incorporating cumulative AG dosage, electrophysiological markers of synaptopathy (e.g., ABR wave I), and multiple SiN paradigms may clarify causal pathways and identify individuals at highest risk for functional decline. The use of monaural, sentence-based SiN may have limited sensitivity to EHF contributions; future work should include tasks with more high-frequency informational weight and spatial listening demands.

## Conclusions

In summary, although EHFHL was more common among individuals with CF exposed to AGs compared to healthy controls, functional SiN deficits were driven primarily by SF cochlear involvement and by neural factors measured by MEMR growth functions. These findings highlight the value of multidimensional auditory assessment in

CF and suggest that both sensory and neural mechanisms contribute to communication challenges in this population. Individuals with SiN difficulties, therefore, may require both sensory aids (hearing aids) as well as training in strategies to address non-sensory factors, such as metalinguistic and compensatory approaches.

## Data Availability

All data produced in the present study are available upon reasonable request to the authors.

